# Outcome Prediction in Aneurysmal Subarachnoid Hemorrhage with World Federation of Neurological Societies grade V(OPAS-V)

**DOI:** 10.1101/2023.06.07.23291115

**Authors:** Shuhei Yamada, Takeo Nishida, Tomofumi Takenaka, Hiroki Yamazaki, Ryota Nakagawa, Masatoshi Takagaki, Yoshihiro Yano, Hajime Nakamura, Shingo Toyota, Toshiyuki Fujinaka, Takuyu Taki, Toshiaki Fujita, Haruhiko Kishima

**Author notes:** **Corresponding author:** Takeo Nishida, MD, PhD Department of Neurosurgery, Osaka University Graduate School of Medicine, 2-2 Yamadaoka, Suita, Osaka, 565-0871 Japan.

## Abstract

**Background:** Aneurysmal subarachnoid hemorrhage (aSAH) with World Federation of Neurological Societies (WFNS) grade V has a high mortality rate and poor prognosis. Some patients with WFNS grade V aSAH have had good outcomes after aggressive treatment; however, outcome predictions based on routine examinations and findings obtained at admission are yet to be reported. This study aimed to develop a decision tree model for predicting outcomes of patients with WFNS grade V aSAH to aid decision-making for treatment strategy.

**Methods:** A multicenter study with retrospective and prospective data collected from 201 (derivation cohort) and 12 (validation cohort) patients with WFNS grade V aSAH, respectively, was conducted. Clinical outcomes were divided into good (Modified Rankin Scale [mRS] score at the time of discharge: 0–2) and poor (mRS score: 3–6) outcomes. A decision tree model was developed for the derivation cohort using the classification and regression tree method with clinical data including laboratory findings; it was named OPAS-V (Outcome Prediction in Aneurysmal Subarachnoid hemorrhage with WFNS grade V). The performance of the model was evaluated by area under the curve (AUC) and overall accuracy in both cohorts.

**Results:** OPAS-V comprised 3 metrics; the percentage of lymphocytes (<49.9% or not), age (>50 yrs or not), and glucose to potassium ratio (≥3.2 or not). The model achieved an AUC of 0.828 (95% confidence interval: 0.712–0.944) and overall accuracy of 0.930. Moreover, the model performed well in the validation cohort with an AUC of 0.700 (95% confidence interval: 0.200–1) and overall accuracy of 0.833.

**Conclusions:** This study developed the first decision tree model for predicting outcomes of patients with WFNS grade V aSAH, based on simple findings obtained at admission. This may aid clinicians in determining treatment strategies for severe conditions such as WFNS grade V aSAH.

## Introduction

Aneurysmal subarachnoid hemorrhage (aSAH) causes severe neurological deficit and mortality, despite recent progress in treatment.^1^ The World Federation of Neurological Societies (WFNS) Scale has been the most significant outcome predictor in several large-scale prognostic studies conducted on patients with aSAH.^2–4^ Recent studies focusing on patients with poor-grade aSAH have also revealed a clear impact of WFNS grade V on their predicted outcomes.^5, 6^

WFNS grade V aSAH accounts for 20–35% of all cases,^7–10^ and has a high mortality rate of 48–63% during hospitalization.^10, 11^ Conversely, several reports have revealed that 14–25% of aggressively treated patients with WFNS grade V aSAH had good outcomes.^9, 11–14^ The clinical severity of aSAH at admission has been reported to reflect the acute inflammatory response, including early brain injury (EBI).^15–17^ In patients with WFNS grade V, the impact of EBI on their outcomes may be heterogeneous^16, 18^ and should be considered when developing outcome prediction models.

Laboratory findings are usually reviewed at admission for aSAH as well as physical and radiological examinations. Many studies have reported that specific blood test findings at admission correlate with EBI or outcomes after aSAH.^17–31^ However, no previous study has successfully predicted the outcome in patients with WFNS grade V aSAH, based on the routine examinations and findings obtained at admission, including these laboratory findings.

This retrospective multicenter study aimed to develop a decision tree model for predicting outcomes of patients with WFNS grade V aSAH using routine assessment findings obtained at admission to aid decision-making for treatment strategy.

## Methods

### Data Availability

The anonymized data that support the findings of this study are available from the corresponding author upon reasonable request.

### Study Design and Patient Selection

We conducted a multicenter study of patients with severe aSAH, defined as grade V, according to the WFNS scale. We collected retrospective data from patients who were admitted to 4 institutions (Osaka University Hospital, Osaka, Japan; Osaka National Hospital, Osaka, Japan; Hanwa Memorial Hospital, Osaka, Japan; and Kansai Rosai Hospital, Hyogo, Japan) between August 2012 and June 2021, for the derivation cohort. We defined the validation cohort as prospective data from patients who were admitted to the same institutions between July 2021 and February 2022. The inclusion criteria were as follows: (1) patients diagnosed with SAH using computed tomography (CT); (2) patients with saccular aneurysms confirmed via three-dimensional CT angiography (CTA) or digital subtraction angiography at admission; (3) patients with WFNS grade V; (4) patients who underwent surgical intervention for aneurysmal occlusion; and (5) patients whose family members provided informed consent for surgical intervention. We excluded patients with a pre-stroke Modified Rankin Scale (mRS) score ≥3.

The Osaka University Clinical Research Review Committee approved the study (approval number 19486) and waived the need for additional written informed consent. This study adhered to the STROBE (Strengthening the Reporting of Observational Studies in Epidemiology) reporting guideline.^32^

### Outcomes

We assessed clinical outcomes using mRS score at the time of discharge. We defined good and poor outcomes as mRS scores of 0–2 and 3–6, respectively.^12, 25^

### Clinical Data Collection

We retrospectively collected patients’ characteristics from the medical records. The lead author (S.Y.) carefully determined radiological findings (for example, Fisher group^33^ or modified Fisher grade^34^) via initial CT, which was independently confirmed by the senior author (T.N.). We collected the initial laboratory findings at admission or within the initial 24 h after admission, and calculated the neutrophil to lymphocyte ratio (NLR),^20, 24, 25^ glucose to potassium ratio,^28^ and C-reactive protein (CRP) to albumin ratio.^31^ We used SI units for the laboratory findings.^35^

### Decision Tree Model Development

We used the classification and regression tree (CART) method as the learning algorithm.^36^ The CART method is nonparametric and does not depend on a specific type of data distribution,^37^; hence, we included all variables in the CART analysis. We substituted missing values with the most frequent value and median value for categorical and continuous variables, respectively. We evaluated the performance of a decision tree model by area under the curve (AUC) of receiver operating characteristic (ROC) analysis, overall accuracy, sensitivity, and specificity. Overall accuracy was the ratio of correctly predicted outcomes. Sensitivity and specificity were the ratios of the predicted good or poor outcomes, respectively, that had actually developed.

### Statistical Analysis

All statistical analyses were performed using R 4.1.1 for Windows (www.R-project.org; R Foundation for Statistical Computing, Vienna, Austria). Categorical variables were assessed using Fisher’s exact test and presented as frequency (percentages). Continuous variables were assessed using the Mann–Whitney U test and presented as median and interquartile range (IQR). Multivariate logistic regression analysis was performed with variables that were significant in the univariate analyses. Statistical significance was set at *p* < 0.05.

## Results

### Patient Characteristics and Outcomes

In the derivation cohort, 210 patients with WFNS grade V aSAH underwent surgical intervention for aneurysmal occlusion. Among them, 9 were excluded because of their poor pre-stroke mRS score ≥3. Thus, the data of 201 (67 male and 134 female) and 12 (1 male and 11 female) patients in the derivation and validation cohorts, respectively, were independently analyzed. Patient characteristics are presented in **Table 1**. The median age at aSAH onset was 70 (IQR: 58–79 years) and 59 (IQR: 55–67 years) years in the derivation and validation cohorts, respectively.

**Table 1.**
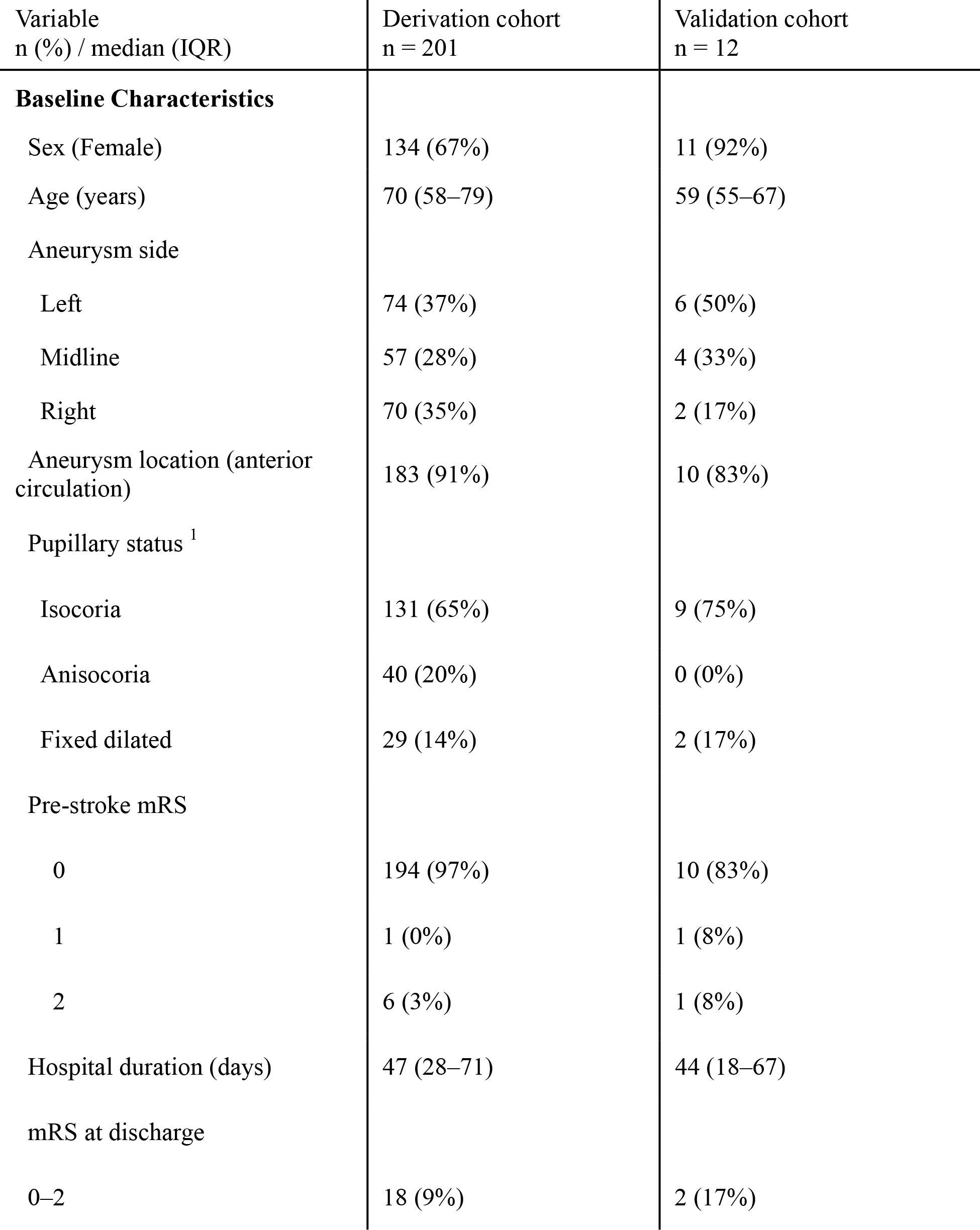
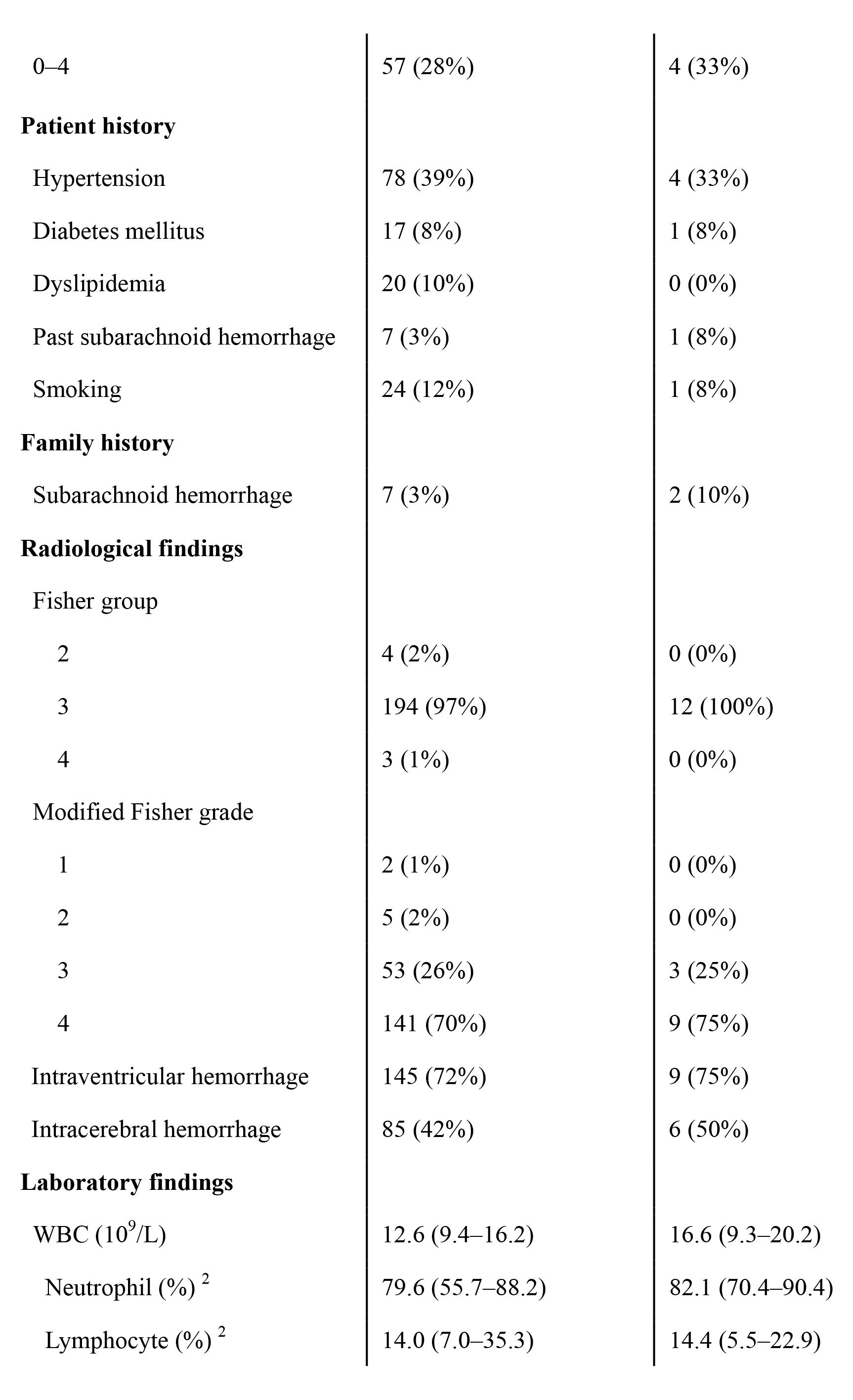
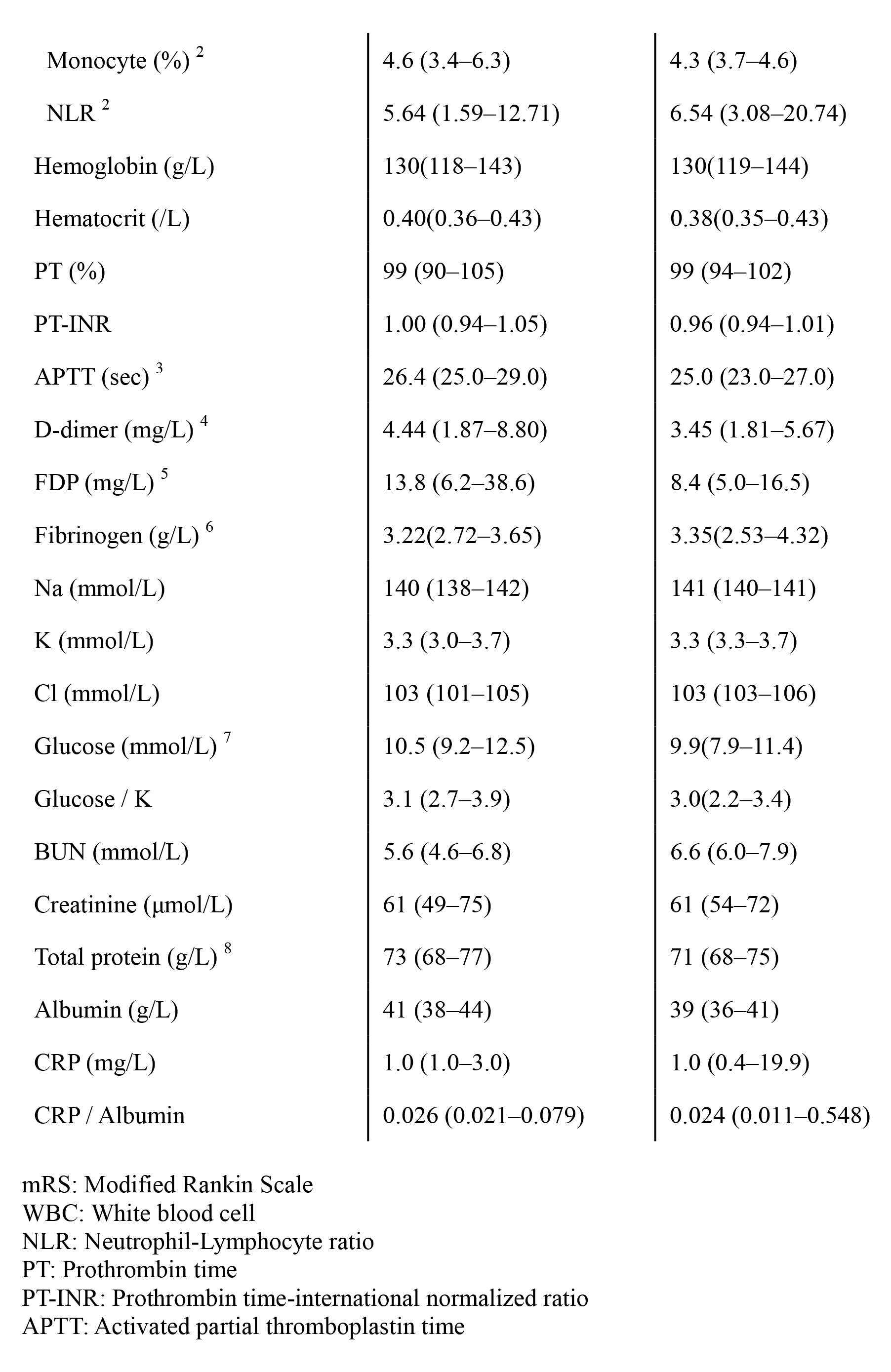
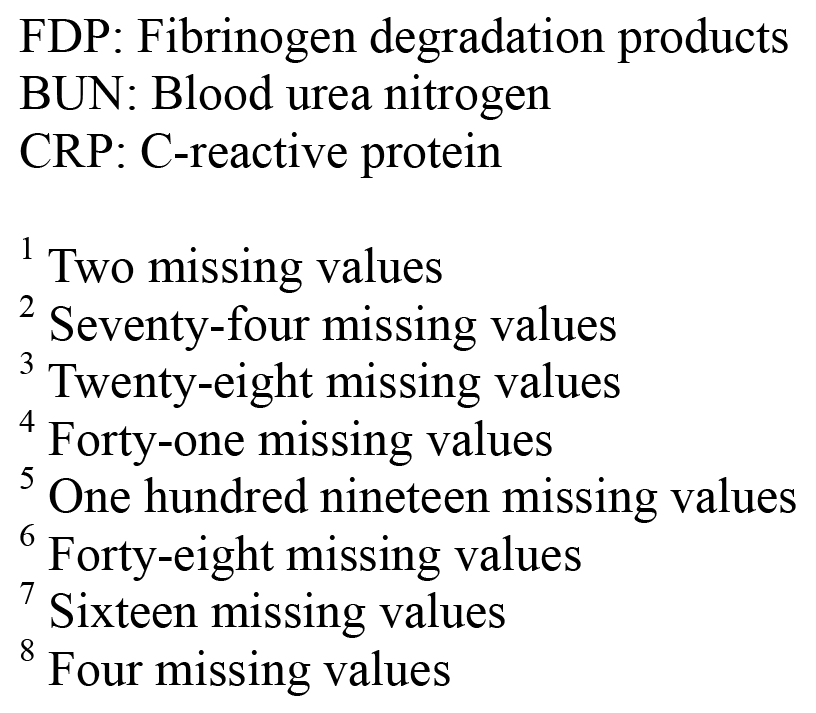
Patient characteristics.

The median duration of hospital stay was 47 (IQR: 28–71 days) and 44 (IQR: 18– 67 days) days in the derivation and validation cohorts, respectively. Eighteen (9%) and 2 (17%) patients in the derivation and validation cohorts, respectively, had good outcomes (mRS score 0–2).

### Decision Tree Model Distinguishing between mRS Score 0–2 and 3–6

We developed a decision tree model, termed OPAS-V (Outcome Prediction in Aneurysmal Subarachnoid hemorrhage with World Federation of Neurological Societies grade V), to distinguish between mRS score 0–2 (good outcome) and 3–6 (poor outcome) using the data of the derivation cohort (**Figure 1**). OPAS-V consisted of 3 metrics; the percentage of lymphocytes, age, and glucose to potassium ratio. If the percentage of lymphocytes was >49.9% at the first node, the patient had a good outcome (50%). If not, the next node was age; the patient had a poor outcome (97%), if >50 years old. If not, the last node was the glucose to potassium ratio. If it was <3.2, the patient had a good outcome (70%), and if not, a poor outcome (93%). OPAS-V achieved an AUC of 0.828 (95% confidence interval [CI]: 0.712–0.944), overall accuracy of 0.930, sensitivity of 0.667, and specificity of 0.956.

**Figure 1.**
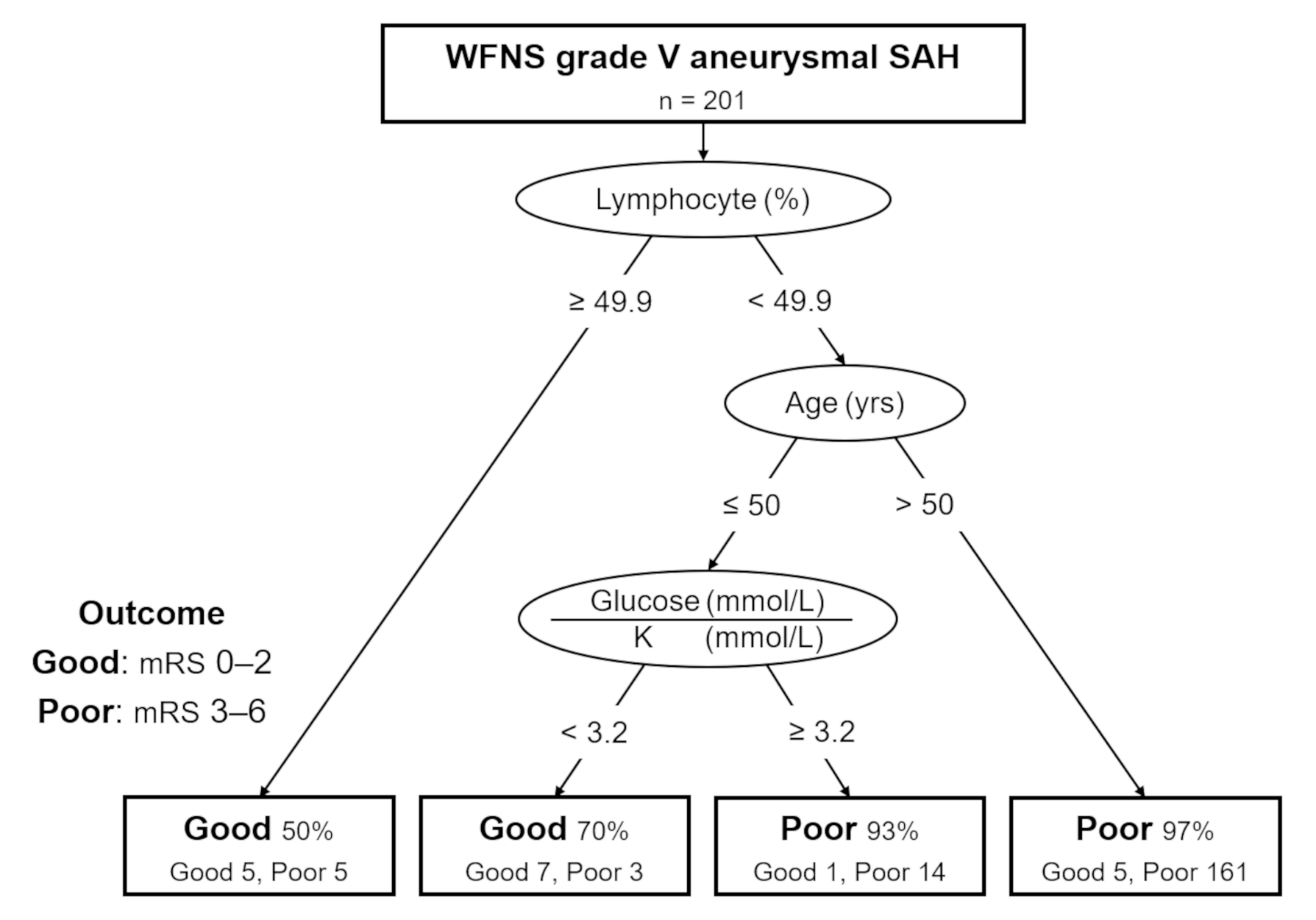
Outcome Prediction in Aneurysmal subarachnoid hemorrhage with World Federation of Neurological Societies grade V (OPAS-V) We have defined good and poor outcomes as modified Rankin Scale scores of 0–2 and 3–6, respectively, at the time of discharge. OPAS-V comprised 3 metrics; the percentage of lymphocytes, age, and glucose to potassium ratio. K: potassium, WFNS: World Federation of Neurological Societies, SAH: subarachnoid hemorrhage

Furthermore, OPAS-V performed well in the validation cohort with an AUC of 0.700 (95% CI: 0.200–1), overall accuracy of 0.833, sensitivity of 0.500, and specificity of 0.900.

### Comparison of Outcomes in the Derivation Cohort

Univariate analysis revealed significant differences between the outcomes (mRS score 0–2 and 3–6) in the derivation cohort in terms of age (*p* < 0.001), Fisher group (*p* = 0.009), modified Fisher grade (*p* = 0.018), intracerebral hemorrhage (ICH) (*p* = 0.005), percentage of neutrophils (*p* = 0.001), percentage of lymphocytes (*p* = 0.002), NLR (*p* = 0.002), and blood urea nitrogen (*p* = 0.042) (**Table 2**). For multivariate analysis, Fisher group was excluded as it is similar to modified Fisher grade. Additionally, the percentage of neutrophils and NLR were excluded as they are related to the percentage of lymphocytes. Multivariate analysis for a poor outcome (mRS score 3–6) revealed significant differences in terms of age (*p* = 0.001, odds ratio [OR]: 1.08 [95% CI: 1.03– 1.14]), ICH (*p* = 0.005, OR: 17.37 [95% CI: 2.39–126.34]), and the percentage of lymphocytes (*p* = 0.013, OR: 0.96 [95% CI: 0.92–0.99]) (**Table 3**).

**Table 2.** Comparison of outcomes in the derivation cohort.

| Variable<br>n (%) / median (IQR) | Good outcome (mRS<br>score: 0–2)<br>n = 18 (9%) | Poor outcome (mRS<br>score: 3–6)<br>n = 183 (91%) | p-value |
| --- | --- | --- | --- |
| **Baseline Characteristics** |  |  |  |
| Sex (Female) | 14 (78%) | 120 (66%) | 0.433 |
| Age (years) | 55 (46–64) | 71 (60–79) | < 0.001 |
| Aneurysm side |  |  | 0.526 |
| Left | 9 (50%) | 65 (36%) |  |
| Midline | 4 (22%) | 53 (29%) |  |
| Right | 5 (28%) | 65 (36%) |  |
| Aneurysm location (anterior<br>circulation) | 16 (89%) | 167 (91%) | 0.667 |
| Pupillary status <sup>1</sup> |  |  | 0.346 |
| Isocoria | 15 (83%) | 116 (63%) |  |
| Anisocoria | 2 (11%) | 38 (21%) |  |
| Fixed dilated | 1 (6%) | 28 (15%) |  |
| Pre-stroke mRS |  |  | 1 |
| 0 | 18 (100%) | 176 (96%) |  |
| 1 | 0 (0%) | 1 (1%) |  |
| 2 | 0 (0%) | 6 (3%) |  |
| **Patient history** |  |  |  |
| Hypertension | 5 (28%) | 73 (40%) | 0.448 |
| Diabetes mellitus | 0 (0%) | 17 (9%) | 0.373 |
| Dyslipidemia | 3 (17%) | 17 (9%) | 0.398 |
| Past subarachnoid hemorrhage | 0 (0%) | 7 (4%) | 1 |
| Smoking | 2 (11%) | 22 (12%) | 1 |
| **Family history** |  |  |  |
| Subarachnoid hemorrhage | 0 (0%) | 7 (4%) | 1 |
| **Radiological findings** |  |  |  |
| Fisher group |  |  | 0.009 |
| 2 | 1 (6%) | 3 (2%) |  |
| 3 | 15 (83%) | 179 (98%) |  |
| 4 | 2 (11%) | 1 (1%) |  |
| Modified Fisher grade |  |  | 0.018 |
| 1 | 1 (6%) | 1 (1%) |  |
| 2 | 2 (11%) | 3 (2%) |  |
| 3 | 6 (33%) | 47 (26%) |  |
| 4 | 9 (50%) | 132 (72%) |  |
| Intraventricular hemorrhage | 10 (56%) | 135 (74%) | 0.107 |
| Intracerebral hemorrhage | 2 (11%) | 83 (45%) | 0.005 |
| **Laboratory findings** |  |  |  |
| WBC (10<sup>9</sup>/L) | 11.8 (9.4–12.9) | 13.0 (9.4–16.3) | 0.171 |
| Neutrophil (%) <sup>2</sup> | 44.9 (38.2–66.5) | 80.9 (61.0–88.3) | 0.001 |
| Lymphocyte (%) <sup>2</sup> | 48.2 (26.2–53.6) | 12.8 (6.6–32.7) | 0.002 |
| Monocyte (%) <sup>2</sup> | 6.2 (4.7–6.7) | 4.6 (3.3–6.1) | 0.124 |
| NLR <sup>2</sup> | 0.93 (0.71–2.78) | 6.46 (1.87–13.33) | 0.002 |
| Hemoglobin (g/L) | 130 (124–133) | 133 (118–143) | 0.537 |
| Hematocrit (/L) | 0.39 (0.38–0.40) | 0.40 (0.36–0.43) | 0.707 |
| PT (%) | 100 (89–106) | 99 (90–105) | 0.892 |
| PT-INR | 0.99 (0.94–1.10) | 1.00 (0.94–1.05) | 0.753 |
| APTT (sec) <sup>3</sup> | 26.3 (25.5–27.7) | 26.6 (25.0–29.0) | 0.967 |
| D-dimer (mg/L) <sup>4</sup> | 2.84 (1.67–6.59) | 4.45 (1.89–8.83) | 0.376 |
| FDP (mg/L) <sup>5</sup> | 7.5 (6.5–18.4) | 14.0 (6.3–39.2) | 0.502 |
| Fibrinogen (g/L) <sup>6</sup> | 3.06 (2.43–3.48) | 3.24 (2.75–3.64) | 0.463 |
| Na (mmol/L) | 140 (138–142) | 140 (138–142) | 0.917 |
| K (mmol/L) | 3.3 (2.9–3.4) | 3.4 (3–3.7) | 0.100 |
| Cl (mmol/L) | 104 (102–105) | 103 (101–105) | 0.533 |
| Glucose (mmol/L) <sup>7</sup> | 9.7 (8.0–11.7) | 10.5 (9.2–12.6) | 0.069 |
| Glucose / K | 3.1 (2.6–3.6) | 3.1 (2.7–4.0) | 0.487 |
| BUN (mmol/L) | 4.8 (4.3–5.7) | 5.7 (4.6–6.8) | 0.042 |
| Creatinine (μmol/L) | 57 (45–65) | 61 (49–77) | 0.218 |
| Total protein (g/L) <sup>8</sup> | 72 (69–75) | 73 (68–77) | 0.811 |
| Albumin (g/L) | 41 (39–43) | 41 (38–44) | 0.900 |
| CRP (mg/L) | 1.0 (1.0–3.0) | 1.0 (0.9–3.0) | 0.740 |
| CRP / Albumin | 0.028 (0.024–0.066) | 0.026 (0.021–0.080) | 0.742 |

mRS: Modified Rankin Scale
WBC: White blood cell
NLR: Neutrophil-Lymphocyte ratio
PT: Prothrombin time
PT-INR: Prothrombin time-international normalized ratio
APTT: Activated partial thromboplastin time
FDP: Fibrinogen degradation products
BUN: Blood urea nitrogen
CRP: C-reactive protein

**Table 3.** Results of multiple logistic regression analysis for a poor outcome (mRS score: 3–6)

| Variable | *p*-value | Odds ratio (95% CI) |
| --- | --- | --- |
| Age | 0.001 | 1.08 (1.03–1.14) |
| Modified Fisher grade |  |  |
| 1 | - | 1.0 (reference) |
| 2 | 0.414 | 0.19 (0.004–9.96) |
| 3 | 0.326 | 5.22 (0.19–141.50) |
| 4 | 0.198 | 8.51 (0.33–221.45) |
| Intracerebral hemorrhage |  |  |
| No | - | 1.0 (reference) |
| Yes | 0.005 | 17.37 (2.39–126.34) |
| Lymphocyte (%) | 0.013 | 0.96 (0.92–0.99) |
| BUN | 0.237 | 1.10 (0.94–1.30) |

mRS: Modified Rankin Scale
CI: Confidence Interval
BUN: Blood urea nitrogen

## Discussion

We have developed a decision tree model comprising simple findings obtained at admission, which accurately predict the outcome of patients with WFNS grade V aSAH and aid in clinical decision-making, including treatment strategies.

Maximal treatment for all patients with WFNS grade V aSAH is challenging and controversial due to high mortality rates and poor prognoses.^10, 13^ Conversely, several reports have revealed that some of patients have good outcomes.^9, 11–14^ It is still unclear which factors develops this heterogeneity in their outcomes. However, our decision tree model, OPAS-V, based on the routine examinations including laboratory findings at admission may be useful in identifying patients who need aggressive treatment. This may be because the laboratory findings included in OPAS-V indicate a certain impact of EBI on the outcome of patients with aSAH. Further external validation is needed because of the small number of patients in the validation cohort.

OPAS-V comprised 3 metrics; the percentage of lymphocytes, age, and glucose to potassium ratio. The nodes derived by CART analysis are not necessarily known predictors. However, age was a correlated metric with a good outcome at the time of discharge,^38^ as confirmed in this study. The glucose to potassium ratio and NLR, which is related to the acute inflammatory response and percentage of lymphocytes, have also frequently been reported to be correlated with aSAH and other acute brain injury-related diseases, in recent years.^17, 20, 24–26, 28, 39–42^ Further studies should focus on laboratory findings, including these indices, in patients with WFNS grade V aSAH.

Few prognostic models have considered laboratory findings obtained at admission, in addition to physical and radiological findings.^23^ Our model is applicable even to WFNS grade V because many clinicians routinely obtain the components of our model during patient assessment at admission. The Glasgow Coma Scale motor value on the second day of hospitalization or CTA source images at admission may impact the outcome of WFNS grade V aSAH.^9, 12^ Furthermore, uncommon laboratory findings such as serum S100 and serum adipocyte fatty acid-binding proteins have been reported to correlate with the prognosis of aSAH.^43–45^ However, these findings might be difficult for application in daily clinical practice.

The SAFIRE grading scale, a recent large-scale study including all WFNS grade patients, reported good discrimination with internal and external AUCs of 0.90 and 0.73, respectively.^2^ However, WFNS grade V has >50% risk of poor outcomes, and only insufficient subdivision has been achieved.^2^ This is partly because clinical course and functional prognosis are significantly correlated with the severity of aSAH.^38, 46, 47^ Further studies focused on patients with WFNS grade V aSAH are required.

A recent report of poor-grade aSAH revealed an AUC of 0.844 and 0.831 in the derivation and validation cohorts, respectively.^5^ However, delayed cerebral ischemia and shunt-dependent hydrocephalus, which are yet to be determined at admission, were included in the prognostic model.^5^ Another study of poor-grade aSAH developed a decision tree model with internal and external AUCs of 0.88 and 0.94, respectively.^6^ However, this decision tree was more complex than OPAS-V and included pupillary reactivity, which was not assessed using the standardized protocol, as mentioned in their limitation.^6^

This study has some limitations. First, it was performed in Japan; hence the treatment strategy for WFNS grade V aSAH may differ from that in other countries.^46–48^ Second, the outcome was set as mRS at the time of discharge. This may lead to overlooking patients who may have favorable outcomes a few years after discharge.^14^ Third, patients who did not undergo surgical intervention for aneurysmal occlusion were not included; hence, their outcomes remain unknown.

In conclusion, we have developed the first decision tree model, OPAS-V, to predict the outcome of patients with WFNS grade V aSAH based on the simple findings obtained at admission. Furthermore, OPAS-V comprised 3 metrics; the percentage of lymphocytes, age, and glucose to potassium ratio. This may help clinicians in determining treatment strategies for severe conditions such as WFNS grade V aSAH, at admission.

## Non-standard Abbreviations and Acronyms

aSAH: aneurysmal subarachnoid hemorrhage
AUC: area under the curve
CART: classification and regression tree
CRP: C-reactive protein
CT: computed tomography
CTA: CT angiography
EBI: early brain injury
ICH: intracerebral hemorrhage
IQR: interquartile range
mRS: modified Rankin Scale
NLR: neutrophil to lymphocyte ratio
OPAS-V: Outcome Prediction in Aneurysmal Subarachnoid hemorrhage with WFNS grade V
ROC: receiver operating characteristic
WFNS: World Federation of Neurological Societies

## Data Availability

All data referred to in the manuscript is available.

## Acknowledgments

We would like to thank Editage (www.editage.com) for English language editing.

## References

1. Macdonald RL, Schweizer TA. Spontaneous subarachnoid haemorrhage. Lancet [Internet]. 2017;389:655–666. Available from: http://dx.doi.org/10.1016/S0140-6736(16)30668-7

2. Van Donkelaar CE, Bakker NA, Birks J, Veeger NJGM, Metzemaekers JDM, Molyneux AJ, Groen RJM, Van Dijk JMC. Prediction of outcome after aneurysmal subarachnoid hemorrhage: Development and validation of the SAFIRE grading scale. Stroke. 2019;50:837–844.

3. Jaja BNR, Saposnik G, Lingsma HF, Macdonald E, Thorpe KE, Mamdani M, Steyerberg EW, Molyneux A, Manoel ALDO, Schatlo B, et al. Development and validation of outcome prediction models for aneurysmal subarachnoid haemorrhage: The SAHIT multinational cohort study. BMJ [Internet]. 2018;360:1–17. Available from: http://dx.doi.org/doi:10.1136/bmj.j5745

4. Risselada R, Lingsma HF, Bauer-Mehren A, Friedrich CM, Molyneux AJ, Kerr RSC, Yarnold J, Sneade M, Steyerberg EW, Sturkenboom MCJM. Prediction of 60 day case-fatality after aneurysmal subarachnoid haemorrhage: Results from the International Subarachnoid Aneurysm Trial (ISAT). Eur. J. Epidemiol. 2010;25:261–266.

5. Shen J, Yu J, Huang S, Mungur R, Huang K, Pan X, Yu G, Xie Z, Zhou L, Liu Z, et al. Scoring model to predict functional outcome in poor-grade aneurysmal subarachnoid hemorrhage. Front. Neurol. 2021;12:1–2.

6. Liu J, Xiong Y, Zhong M, Yang Y, Guo X, Tan X, Zhao B. Predicting long-term outcomes after poor-grade aneurysmal subarachnoid hemorrhage using decision tree modeling. Neurosurgery. 2020;87:523–529.

7. Udy AA, Vladic C, Saxby ER, Cohen J, Delaney A, Flower O, Anstey M, Bellomo R, Cooper DJ, Pilcher D V. Subarachnoid hemorrhage patients admitted to intensive care in Australia and New Zealand: A multicenter cohort analysis of in-hospital mortality over 15 years. Crit. Care Med. 2017;45:e135–e145.

8. Van Heuven AW, Mees SMD, Algra A, Rinkel GJE. Validation of a prognostic subarachnoid hemorrhage grading scale derived directly from the glasgow coma scale. Stroke. 2008;39:1347–1348.

9. Van Den Berg R, Foumani M, Schröder RD, Peerdeman SM, Horn J, Bipat S, Vandertop WP. Predictors of outcome in World Federation of Neurologic Surgeons grade v aneurysmal subarachnoid hemorrhage patients. Crit. Care Med. 2011;39:2722–2727.

10. Ojha M, Finnis ME, Heckelmann M, Raith EP, Moodie S, Chapman MJ, Reddi B, Maiden MJ. Outcomes following grade V subarachnoid haemorrhage: A single-centre retrospective study. Anaesth. Intensive Care. 2020;48:289–296.

11. Wostrack M, Sandow N, Vajkoczy P, Schatlo B, Bijlenga P, Schaller K, Kehl V, Harmening K, Ringel F, Ryang YM, et al. Subarachnoid haemorrhage WFNS grade V: Is maximal treatment worthwhile? Acta Neurochir. (Wien). 2013;155:579– 586.

12. Sorimachi T, Osada T, Aoki R, Nishiyama J, Hirayama A, Srivatanakul K, Matsumae M. Density of the cerebral cortex in computed tomography angiography source images and clinical outcomes in Grade V subarachnoid hemorrhage. Neurol. Res. 2015;37:484–490.

13. Kijima N, Nakagawa T, Miura S, Nakagawa R, Tachi T, Okita Y, Kanemura Y, Nakajima S, Fujinaka T. Therapeutic strategies for poor-grade subarachnoid hemorrhage patients and clinical outcomes. Surg. Cereb. Stroke [Internet]. 2021;49:98–102. Available from: https://www.jstage.jst.go.jp/article/scs/49/2/49_98/_article/-char/ja/

14. Ariyada K, Ohida T, Shibahashi K, Hoda H, Hanakawa K, Murao M. Long-term functional outcomes for world federation of neurosurgical societies grade v aneurysmal subarachnoid hemorrhage after active treatment. Neurol. Med. Chir. (Tokyo). 2020;60:390–396.

15. Hanafy KA, Morgan Stuart R, Fernandez L, Schmidt JM, Claassen J, Lee K, Sander Connolly E, Mayer SA, Badjatia N. Cerebral inflammatory response and predictors of admission clinical grade after aneurysmal subarachnoid hemorrhage. J. Clin. Neurosci. 2010;17:22–25.

16. Rass V, Helbok R. Early Brain Injury After Poor-Grade Subarachnoid Hemorrhage. Curr. Neurol. Neurosci. Rep. 2019;19.

17. Kawabata S, Takagaki M, Nakamura H, Nishida T, Terada E, Kadono Y, Izutsu N, Takenaka T, Matsui Y, Yamada S, et al. Association of Gut Microbiome with Early Brain Injury After Subarachnoid Hemorrhage: an Experimental Study. Transl. Stroke Res. [Internet]. 2022;Available from: https://doi.org/10.1007/s12975-022-01112-6

18. Takahashi S, Akiyama T, Horiguchi T, Miwa T, Takemura R, Yoshida K. Loss of consciousness at ictus and/or poor World Federation of Neurosurgical Societies grade on admission reflects the impact of EBI and predicts poor outcome in patients with SAH. Surg. Neurol. Int. 2020;11.

19. Krzyżewski RM, Kliś KM, Kwinta BM, Stachura K, Guzik TJ, Gąsowski J. High leukocyte count and risk of poor outcome after subarachnoid hemorrhage: A meta-analysis. World Neurosurg. 2020;135:e541–e547.

20. Tao C, Wang J, Hu X, Ma J, Li H, You C. Clinical value of neutrophil to lymphocyte and platelet to lymphocyte ratio after aneurysmal subarachnoid hemorrhage. Neurocrit. Care. 2017;26:393–401.

21. Liu JH, Li XK, Chen ZB, Cai Q, Wang L, Ye YH, Chen QX. D-dimer may predict poor outcomes in patients with aneurysmal subarachnoid hemorrhage: A retrospective study. Neural Regen. Res. 2017;12:2014–2020.

22. Xie B, Lin Y, Wu X, Yu L, Zheng S, Kang D. Reduced admission serum fibrinogen levels predict 6-month mortality of poor-grade aneurysmal subarachnoid hemorrhage. World Neurosurg. [Internet]. 2020;136:e24–e32. Available from: https://doi.org/10.1016/j.wneu.2019.08.155

23. Li R, Lin F, Chen Y, Lu J, Han H, Ma L, Zhao Y, Yan D, Li R, Yang J, et al. A 90-Day Prognostic Model Based on the Early Brain Injury Indicators after Aneurysmal Subarachnoid Hemorrhage: the TAPS Score. Transl. Stroke Res. 2022;

24. Giede-Jeppe A, Reichl J, Sprügel MI, Lücking H, Hoelter P, Eyüpoglu IY, Kuramatsu JB, Huttner HB, Gerner ST. Neutrophil-to-lymphocyte ratio as an independent predictor for unfavorable functional outcome in aneurysmal subarachnoid hemorrhage. J. Neurosurg. 2020;132:400–407.

25. Chang JJ, Dowlati E, Triano M, Kalegha E, Krishnan R, Kasturiarachi BM, Gachechiladze L, Pandhi A, Themistocleous M, Katsanos AH, et al. Admission neutrophil to lymphocyte ratio for predicting outcome in subarachnoid hemorrhage. J. Stroke Cerebrovasc. Dis. [Internet]. 2021;30:105936. Available from: https://doi.org/10.1016/j.jstrokecerebrovasdis.2021.105936

26. Nóbrega Lima Rodrigues de Morais A, Ribeiro Baylão VM, Martins Silva T, Gomes dos Santos A, Azevedo M J. M. de Oliveira A. Is neutrophil-lymphocyte ratio a useful tool for predicting outcome in subarachnoid hemorrhage? A systematic review. Neurosurg. Rev. 2021;44:3023–3028.

27. Mocco J, Ransom ER, Komotar RJ, Schmidt JM, Sciacca RR, Mayer SA, Connolly ES. Preoperative prediction of long-term outcome in poor-grade aneurysmal subarachnoid hemorrhage. Neurosurgery. 2006;59:529–537.

28. Fujiki Y, Matano F, Mizunari T, Murai Y, Tateyama K, Koketsu K, Kubota A, Kobayashi S, Yokota H, Morita A. Serum glucose/potassium ratio as a clinical risk factor for aneurysmal subarachnoid hemorrhage. J. Neurosurg. 2018;129:870– 875.

29. Romero FR, Bertolini E de F, Figueiredo EG, Teixeira MJ. Serum C-reactive protein levels predict neurological outcome after aneurysmal subarachnoid hemorrhage. Arq. Neuropsiquiatr. 2012;70:202–205.

30. Turner CL, Budohoski K, Smith C, Hutchinson PJ, Kirkpatrick PJ. Elevated baseline C-reactive protein as a predictor of outcome after aneurysmal subarachnoid hemorrhage: Data from the Simvastatin in Aneurysmal Subarachnoid Hemorrhage (STASH) trial. Neurosurgery. 2015;77:786–792.

31. Zhang D, Yan H, Wei Y, Liu X, Zhuang Z, Dai W, Li J, Li W, Hang C. C-reactive protein/albumin ratio correlates with disease severity and predicts outcome in patients with aneurysmal subarachnoid hemorrhage. Front. Neurol. 2019;10:1–6.

32. von Elm E, Altman DG, Egger M, Pocock SJ, Gøtzsche PC, Vandenbroucke JP. The Strengthening the Reporting of Observational Studies in Epidemiology (STROBE) statement: guidelines for reporting observational studies. Lancet [Internet]. 2007;370:1453–1457. Available from: https://linkinghub.elsevier.com/retrieve/pii/S014067360761602X

33. Fisher CM, Kistler JP, Davis JM. Relation of cerebral vasospasm to subarachnoid hemorrhage visualized by Computerized Tomographic scanning. Neurosurgery [Internet]. 1980;6:1–9. Available from: https://academic.oup.com/neurosurgery/article-lookup/doi/10.1227/00006123-198001000-00001

34. Frontera JA, Claassen J, Schmidt JM, Wartenberg KE, Temes R, Connolly ES, MacDonald RL, Mayer SA. Prediction of symptomatic vasospasm after subarachnoid hemorrhage: The modified fisher scale. Neurosurgery. 2006;59:21– 26.

35. Young DS. Implementation of SI units for clinical laboratory data. Ann. Intern. Med. 1987;106:114.

36. Breiman L, Friedman JH, Olshen RA, Stone CJ. Classification and regression trees [Internet]. Routledge; 1984. Available from: https://www.taylorfrancis.com/books/9781351460491

37. Ghiasi MM, Zendehboudi S, Mohsenipour AA. Decision tree-based diagnosis of coronary artery disease: CART model. Comput. Methods Programs Biomed. [Internet]. 2020;192:105400. Available from: https://doi.org/10.1016/j.cmpb.2020.105400

38. Ikawa F, Ichihara N, Uno M, Shiokawa Y, Toyoda K, Minematsu K, Kobayashi S, Yamaguchi S, Kurisu K. Visualisation of the non-linear correlation between age and poor outcome in patients with aneurysmal subarachnoid haemorrhage. J. Neurol. Neurosurg. Psychiatry. 2021;92:1173–1180.

39. Zhou J, Yang CS, Shen LJ, Lv QW, Xu QC. Usefulness of serum glucose and potassium ratio as a predictor for 30-day death among patients with severe traumatic brain injury. Clin. Chim. Acta [Internet]. 2020;506:166–171. Available from: https://doi.org/10.1016/j.cca.2020.03.039

40. Wu XY, Zhuang YK, Cai Y, Dong XQ, Wang KY, Du Q, Yu WH. Serum glucose and potassium ratio as a predictive factor for prognosis of acute intracerebral hemorrhage. J. Int. Med. Res. 2021;49.

41. Sabouri E, Majdi A, Jangjui P, Rahigh Aghsan S, Naseri Alavi SA, Siwicka-Gieroba D, Malodobry K, Biernawska J, Robba C, Bohatyrewicz R, et al. The neutrophil/lymphocyte count ratio predicts mortality in severe traumatic brain injury patients. J. Clin. Med. 2019;140:142–147.

42. Lattanzi S, Cagnetti C, Rinaldi C, Angelocola S, Provinciali L, Silvestrini M. Neutrophil-to-lymphocyte ratio improves outcome prediction of acute intracerebral hemorrhage. J. Neurol. Sci. [Internet]. 2018;387:98–102. Available from: https://doi.org/10.1016/j.jns.2018.01.038

43. Kedziora J, Burzynska M, Gozdzik W, Kübler A, Kobylinska K, Adamik B. Biomarkers of neurological outcome after aneurysmal subarachnoid hemorrhage as early predictors at discharge from an intensive care unit. Neurocrit. Care [Internet]. 2021;34:856–866. Available from: https://doi.org/10.1007/s12028-020-01110-2

44. Luo YG, Han B, Sun TW, Liu X, Liu J, Zhang J. The association between serum adipocyte fatty acid-binding protein and 3-month disability outcome after aneurysmal subarachnoid hemorrhage. J. Neuroinflammation. 2020;17:1–12.

45. Shim JH, Yoon SM, Bae HG, Yun IG, Shim JJ, Lee KS, Doh JW. Which treatment modality is more injurious to the brain in patients with subarachnoid hemorrhage? Degree of brain damage assessed by serum S100 protein after aneurysm clipping or coiling. Cerebrovasc. Dis. 2012;34:38–47.

46. Steiner T, Juvela S, Unterberg A, Jung C, Forsting M, Rinkel G. European Stroke Organization guidelines for the management of intracranial aneurysms and subarachnoid haemorrhage. Cerebrovasc. Dis. 2013;35:93–112.

47. Connolly ES, Rabinstein AA, Carhuapoma JR, Derdeyn CP, Dion J, Higashida RT, Hoh BL, Kirkness CJ, Naidech AM, Ogilvy CS, et al. Guidelines for the management of aneurysmal subarachnoid hemorrhage: A guideline for healthcare professionals from the American Heart Association/American Stroke Association. Stroke. 2012;43:1711–1737.

48. Committee for Guidelines for Management of Aneurysmal Subarachnoid, Hemorrhage, Japanese Society on Surgery for Cerebral Stroke. Evidence-based guidelines for the management of aneurysmal subarachnoid hemorrhage. English Edition. Neurol. Med. Chir. (Tokyo). 2012;52:355–429.

